# ClinGen Hereditary Cardiovascular Disease Gene Curation Expert Panel: Reappraisal of Genes associated with Hypertrophic Cardiomyopathy

**DOI:** 10.1101/2024.07.29.24311195

**Authors:** Sophie Hespe, Amber Waddell, Babken Asatryan, Emma Owens, Courtney Thaxton, Mhy-Lanie Adduru, Kailyn Anderson, Emily E. Brown, Lily Hoffman-Andrews, Elizabeth Jordan, Katherine Josephs, Megan Mayers, Stacey Peters, Fergus Stafford, Richard D. Bagnall, Lucas Bronicki, Bert Callewaert, C. Anwar A. Chahal, Cynthia A. James, Olga Jarinova, Andrew P. Landstrom, Elizabeth M. McNally, Brittney Murray, Laura Muiño-Mosquera, Victoria Parikh, Chloe Reuter, Roddy Walsh, Bess Wayburn, James S. Ware, Jodie Ingles

## Abstract

**Background:** Hypertrophic cardiomyopathy (HCM) is an inherited cardiac condition affecting ∼1 in 500 and exhibits marked genetic heterogeneity. Previously published in 2019, 57 HCM-associated genes were curated providing the first systematic evaluation of gene-disease validity. Here we report work by the ClinGen Hereditary Cardiovascular Disorders Gene Curation Expert Panel (HCVD-GCEP) to reappraise the clinical validity of previously curated and new putative HCM genes.

**Methods:** The ClinGen systematic gene curation framework was used to re-classify the gene-disease relationships for HCM and related syndromic entities involving left ventricular hypertrophy. Genes previously curated were included if their classification was not definitive, and if the time since curation was >2-3 years. New genes with literature assertions for HCM were included for initial evaluation. Existing genes were curated for new inheritance patterns where evidence existed. Curations were presented on twice monthly calls, with the HCVD-GCEP composed of 29 individuals from 21 institutions across 6 countries.

**Results:** Thirty-one genes were re-curated and an additional 5 new potential HCM-associated genes were curated. Among the re-curated genes, 17 (55%) genes changed classification: 1 limited and 4 disputed (from no known disease relationship), 9 disputed (from limited), and 3 definitive (from moderate). Among these, 3 (10%) genes had a clinically relevant upgrade, including *TNNC1,* a 9th sarcomere gene with definitive HCM association. With new evidence, two genes were curated for multiple inheritance patterns (*TRIM63,* disputed for autosomal dominant but moderate for autosomal recessive; *ALPK3,* strong for autosomal dominant and definitive for recessive). *CSRP3* was curated for a semi-dominant mode of inheritance (definitive). Nine (29%) genes were downgraded to disputed, further discouraging clinical reporting of variants in these genes. Five genes recently reported to cause HCM were curated: *RPS6KB1* and *RBM20* (limited), *KLHL24* and *MT-TI* (moderate), and *FHOD3* (definitive).

**Conclusions:** We report 29 genes with definitive, strong or moderate evidence of causation for HCM or isolated LVH, including sarcomere, sarcomere-associated and syndromic conditions.

## INTRODUCTION

Hypertrophic cardiomyopathy (HCM), is characterized by left ventricular hypertrophy (LVH) in the absence of abnormal loading conditions.^1,2^ HCM is an inherited cardiomyopathy affecting ∼1 in 500 in the general population,^3^ with a broad spectrum of clinical outcomes ranging from mild or no symptoms, to heart failure and sudden cardiac death. Clinical genetic testing is a key aspect of clinical care for patients with HCM, and a Class 1 recommendation in all recent guidelines, though guidance on the approach to this lacks consistency.^1,2,4,5^ There is evidence of the role of genetic testing in clarifying the overall diagnosis for patients,^6^ and it is anticipated to play an increasing role in prognostication and therapeutic stratification, but primarily it enables cascade genetic testing of at-risk family members. Genotype-positive relatives are recommended to undergo cardiac screening and surveillance, while genotype-negative members are released from this recommendation.^7^ For this reason, genetic testing for HCM has been established as cost-effective in addition to clinical cardiac screening.^8,9^ The challenges associated with genetic testing are well described, with limitations in our understanding of the genetic architecture of HCM,^10^ lack of ancestral diversity in research and population genomic databases impacting our ability to accurately interpret genetic variants^11,12^ and inappropriate test selection or variant interpretation.^13–15^

Our previous work systematically evaluated 57 HCM genes for the quantity and quality of clinical genetic and experimental data using a scoring matrix and gave a final overall summary classification,^16^ showing definitive relationships for 8 sarcomere genes and 14 other genes associated with diseases that could present as isolated LVH. Importantly we showed two-thirds of previously asserted HCM-associated genes had limited or no evidence of disease relationship. The outcomes of this expert-curated HCM gene list have guided genetic testing and variant interpretation worldwide, contributing to published guidelines and expert consensus statements. Indeed, recent AHA/ACC Guidelines for Management of HCM state 8 sarcomere genes should be tested, and other genes only on suspicion of additional clinical findings to suggest a genocopy.^1^

The ClinGen Hereditary Cardiovascular Disorders (HCVD) Gene Curation Expert Panel comprises experts and curators from 6 countries and is tasked with ongoing curation efforts of cardiovascular diseases not already overseen by a dedicated gene curation expert panel. Here we report the systematic re-appraisal of evidence supporting 31 genes previously curated for their association with HCM, and 5 new HCM-associated genes, using the ClinGen framework for evaluating the clinical validity of gene-disease relationships.^13^

## METHODS

### Personnel

Gene curations were performed and assessed by individuals in the ClinGen HCVD Gene Curation Expert Panel who had various areas of expertise in HCM and the ClinGen gene-disease relationship validity curation framework. This group comprised 29 individuals from 21 institutions across 6 countries (Australia, Belgium, Canada, Netherlands, UK, and USA). This expert panel is a subgroup of the ClinGen Cardiovascular Clinical Domain Working Group (https://clinicalgenome.org/working-groups/clinical-domain/cardiovascular/).

### Criteria for re-curation of genes

Per ClinGen’s gene re-curation policies, genes that were classified as disputed or limited more than 3 years ago, moderate 2 years ago, or strong 3 years ago from the original discovery publication date, were re-curated. Gene-disease relationships that previously reached a refuted, no known disease relationship, or definitive classification were only re-evaluated if new or contradictory evidence had emerged. Re-curation was considered for genes with new disease entity assertions, or inheritance modes, that had been made since the last curation. New genes asserted to cause HCM were identified by members of the group based on expert-opinion and literature review, and underwent the pre-curation process to determine the most appropriate disease entity for the curation.

### Gene classifications

As per ACMG guidance (ref Bean *et al*), gene-disease validity was classified as definitive, strong and moderate, though it should be noted that gene-level evidence in moderate genes is considered emerging and variants are unlikely to be classified above likely pathogenic. Genes of uncertain significance are those with classifications of limited, no evidence or disputed. Genes with disputed evidence are not appropriate for diagnostic reporting for the disease-entity in question, including variants of uncertain significance.

### Curated phenotypes

Previously, the HCM Gene Curation Expert Panel curated 57 genes in total and categorized them as either HCM genes, i.e., where LVH is seen in isolation, or as syndromic genes, where LVH is a phenotypic feature of the syndrome.^16^ Our re-curation effort focused only on syndromic genes where disease presentation could plausibly be confused with HCM. Syndromes were further categorized as intrinsic cardiomyopathy if additional phenotypic features were largely confined to the heart and could not be classified according to existing cardiomyopathy phenotypes. Pre-curation was performed for genes where there was question regarding the best disease entity or inheritance pattern to curate for, as per the ClinGen Lumping and Splitting criteria,^17^ which takes into consideration the disease assertions, molecular mechanisms, phenotypic variability, and an inheritance pattern for the asserted disease entities.

### Gene curation process

Curators completed pre-curations within the ClinGen Gene Tracker,^18^ when necessary, for genes with multiple disease assertions. Once the HCVD Gene Curation Expert Panel determined the appropriate disease entity for curation, curators performed a literature review to gather genetic and experimental evidence pertaining to the gene-disease relationship. Curators used the ClinGen Gene Clinical Validity Curation Process, Standard Operating Procedure version 9 (Supplementary Material) to score the evidence and reach a provisional classification within the ClinGen Gene Curation Interface.^18^ Disease-specific refinements to these rules were developed by the HCVD Gene Curation Expert Panel as needed (Supplementary Material). The HCVD Gene Curation Expert Panel met on a twice monthly call to discuss the curations and reach consensus on final classifications.

All data including pre-curations, final classifications, curation details, and expert panel membership has been made publicly available on the ClinGen website: https://search.clinicalgenome.org/kb/affiliate/10104. No institutional review board approval was required.

## RESULTS

### Gene classifications

Out of the 57 genes previously curated by the HCM Gene Curation Expert Panel,^16^ 30 were selected for re-curation with previous classifications: 4 definitive, 1 strong, 3 moderate, 16 limited, and 7 no-known disease relationship (Table 1). The remainder retained their prior classification (definitive or no-known disease relationship), or were considered to cause LVH only in the presence of other overt syndromic features and were not re-curated. Five additional genes not previously classified were selected for curation. More information and supporting references can be found in the publicly accessible gene summaries (https://search.clinicalgenome.org/kb/affiliate/10104; Supplementary material).

**TABLE 1:** Points and classifications for all genes curated or re-curated for hypertrophic cardiomyopathy.

| Gene | Genetic Evidence (0-12) | Experimental Evidence (0-6) | Total Points | Classification |
| --- | --- | --- | --- | --- |
| *PRKAG2* | 12 | 6 | 18 | Definitive |
| *ACTN2* | 12 | 5.5 | 17.5 | Definitive |
| *CSRP3* | 12 | 5.5 | 17.5 | Definitive |
| *CACNA1C* | 12 | 5 | 17 | Definitive |
| *ALPK3 (AR)* | 12 | 3.5 | 15.5 | Definitive |
| *FHOD3* | 10.9 | 4 | 14.9 | Definitive |
| *FLNC (missense)* | 8.1 | 6 | 14.1 | Definitive |
| *TNNC1* | 5.8 | 6 | 11.8 | Definitive |
| *ALPK3 (AD)* | 12 | 6 | 18 | Strong |
| *TRIM63 (AR)* | 7.2 | 2.5 | 9.7 | Moderate |
| *JPH2* | 3 | 5 | 8 | Moderate |
| *KLHL24* | 6.5 | 0.5 | 7 | Moderate |
| *MT-TI* | 5.5 | 1 | 6.5 | Moderate |
| *TTN* | 1.2 | 5.5 | 6.7 | Limited |
| *RYR2* | 0.9 | 4 | 4.9 | Limited |
| *OBSCN* | 3 | 1 | 4 | Limited |
| *KLF10* | 0.6 | 2.5 | 3.1 | Limited |
| *RPS6KB1* | 0.4 | 2.5 | 2.9 | Limited |
| *PDLIM3* | 0 | 2 | 2 | Limited |
| *NEXN* | 0.5 | 1 | 1.5 | Limited |
| *RBM20* | 1.3 | 0 | 1.3 | Limited |
| *TMPO* | 0.1 | 0 | 0.1 | Limited |
| *CACNB2* | 0 | 3.5 | 3.5 | Disputed |
| *MYOZ2* | 1 | 2.5 | 3.5 | Disputed |
| *TRIM63 (AD)* | 0.85 | 2 | 2.85 | Disputed |
| *TCAP* | 0.4 | 1.5 | 1.9 | Disputed |
| *ANKRD1* | 0.2 | 1.5 | 1.7 | Disputed |
| *CASQ2* | 0.1 | 1.5 | 1.6 | Disputed |
| *MYLK2* | 0 | 1.5 | 1.5 | Disputed |
| *MYPN* | 0 | 1.5 | 1.5 | Disputed |
| *MYH6* | 0.4 | 1 | 1.4 | Disputed |
| **MYOM1** | 0.1 | 0.5 | 0.6 | Disputed |
| **VCL** | 0 | 0.5 | 0.5 | Disputed |
| **CALR3** | 0 | 0 | 0 | Disputed |
| **DSP** | 0 | 0 | 0 | Disputed |
| **KCNQ1** | 0 | 0 | 0 | Disputed |
| **TNNC2** | 0 | 0 | 0 | NKDR |

Bold text indicates genes that had a change in classification. NKDR=No Known Disease Relationship.

A total of 35 genes, curated for 37 entities, underwent curation with the final classifications including 8 definitive, 1 strong, 4 moderate, 9 limited evidence, 1 no known disease relationship, and 14 disputed genes (Table 1). Of the 30 genes re-curated, 17 (57%) had a change of classification; 3 became definitive (from moderate; *ACTN2, CSRP3, TNNC1*), 1 became limited (from no evidence; *TMPO*), 4 disputed (from no evidence; *CACNB2, CASQ2, DSP, KCNQ1*), and 9 disputed (from limited; *ANKRD1*, *CALR3, MYH6, MYLK2, MYOM1, MYOZ2, MYPN, TCAP, VCL*). Three (12%) upgraded gene-disease classifications (moderate to definitive) mean variants can now be classified as likely pathogenic or above, while downgraded gene-disease classification from limited to disputed (9; 30%) or no evidence to limited (1; 3%) alters advice regarding reporting of variants on genetic testing reports for HCM. There were 11 genes that retained their original classification: 1 no evidence (*TNNC2*), 6 limited (*KLF10, NEXN, OBSCN, PDLIM3, RYR2, TTN*), 1 moderate (*JPH2*), and 3 definitive evidence (*CACNA1C, FLNC, PRKAG2*). Following pre-curation discussions, two genes were re-curated for both autosomal dominant and recessive modes of inheritance of HCM, including *TRIM63,* which was classified as disputed and moderate for each mode of inheritance, respectively; and *ALPK3,* which was classified as strong and definitive, respectively. *CSRP*3 was curated for a semi-dominant inheritance mode. Five new genes were selected for initial curation; 2 were adjudicated as limited (*RPS6KB1*, *RBM20*), 2 moderate (*KLHL24*, *MT-TI*), and 1 definitive evidence (*FHOD3*) (Figure 1).

**FIGURE 1:**
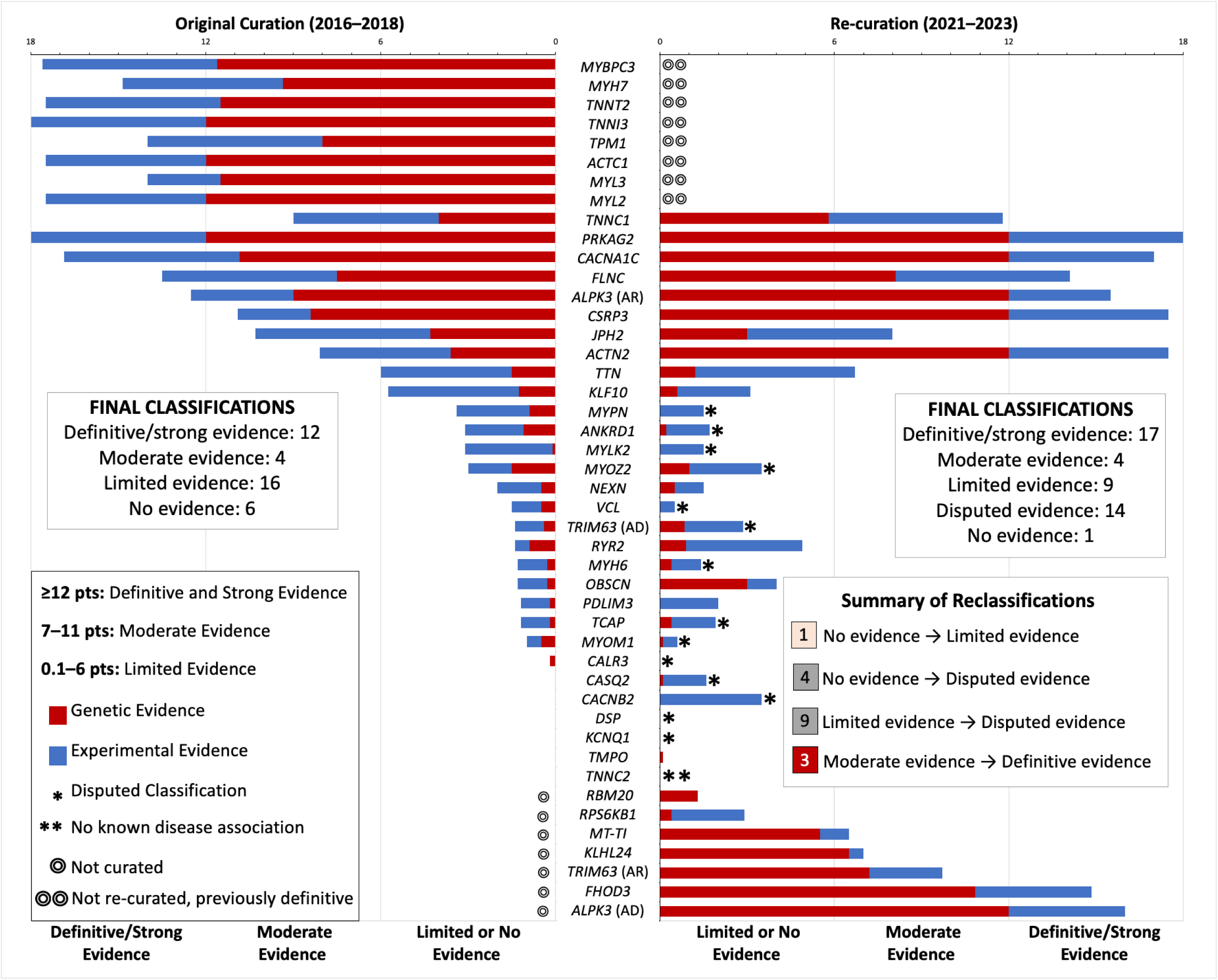
Comparison of original and updated classifications. Hypertrophic cardiomyopathy (HCM) gene-disease classifications showing the genetic and experimental evidence point totals from the original curation compared to the re-curation. Including 17 changes of classification; 3 upgraded from moderate to definitive, 1 upgraded from no evidence to limited, and 13 downgraded to disputed. There are 7 new curations; 5 new genes and 2 existing genes with new modes of inheritance.

### Genes with robust evidence for HCM-association

Overall, there were 29 genes associated with disease, including moderate, strong or definitive levels of evidence, that should be included in HCM genetic testing, and for these we curated inheritance mode, variant classes and mechanism (Table 2; Figure 2). Upgraded gene classifications (definitive: *ACTN2*, *CSRP3*, *TNNC1*) were due to numerous factors, including an increase in genetic evidence derived from literature, including those reporting HCM patient cohorts from geographic regions not typically represented; reconsideration of the inheritance mode or the encompassing phenotype. Further, three new genes achieved classifications that would warrant inclusion in routine HCM genetic testing. Full gene summaries and their associated references are available online (https://search.clinicalgenome.org/kb/gene-validity) and Supplementary Material.

**FIGURE 2:**
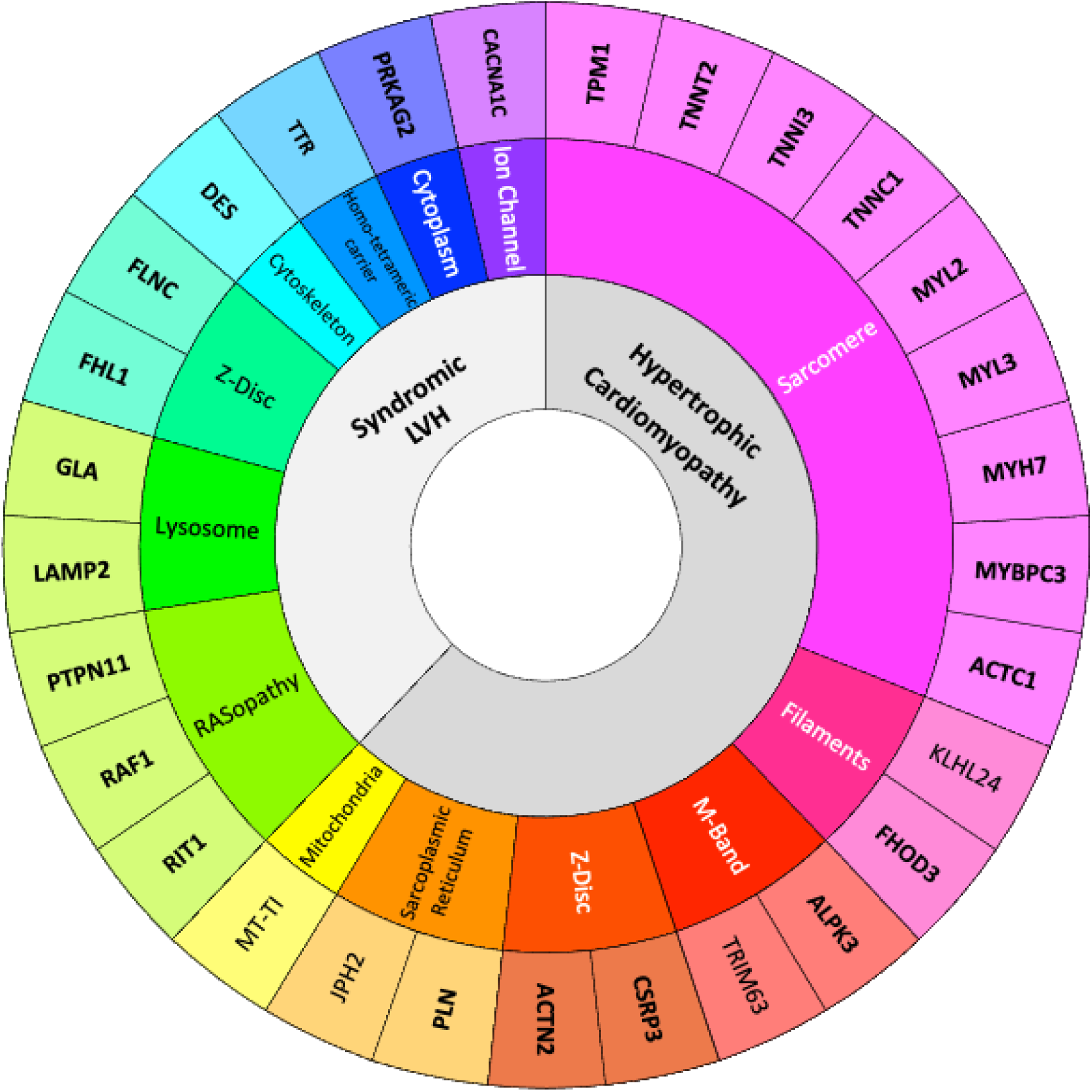
Updated list of genes with moderate, strong or definitive hypertrophic cardiomyopathy (HCM) association. We highlight the genetic architecture of HCM, or genocopies causing left ventricular hypertrophy (innermost circle), spanning numerous gene ontologies (middle circle). Genes classified as definitive or strong evidence are emphasized in bold.

**TABLE 2:**
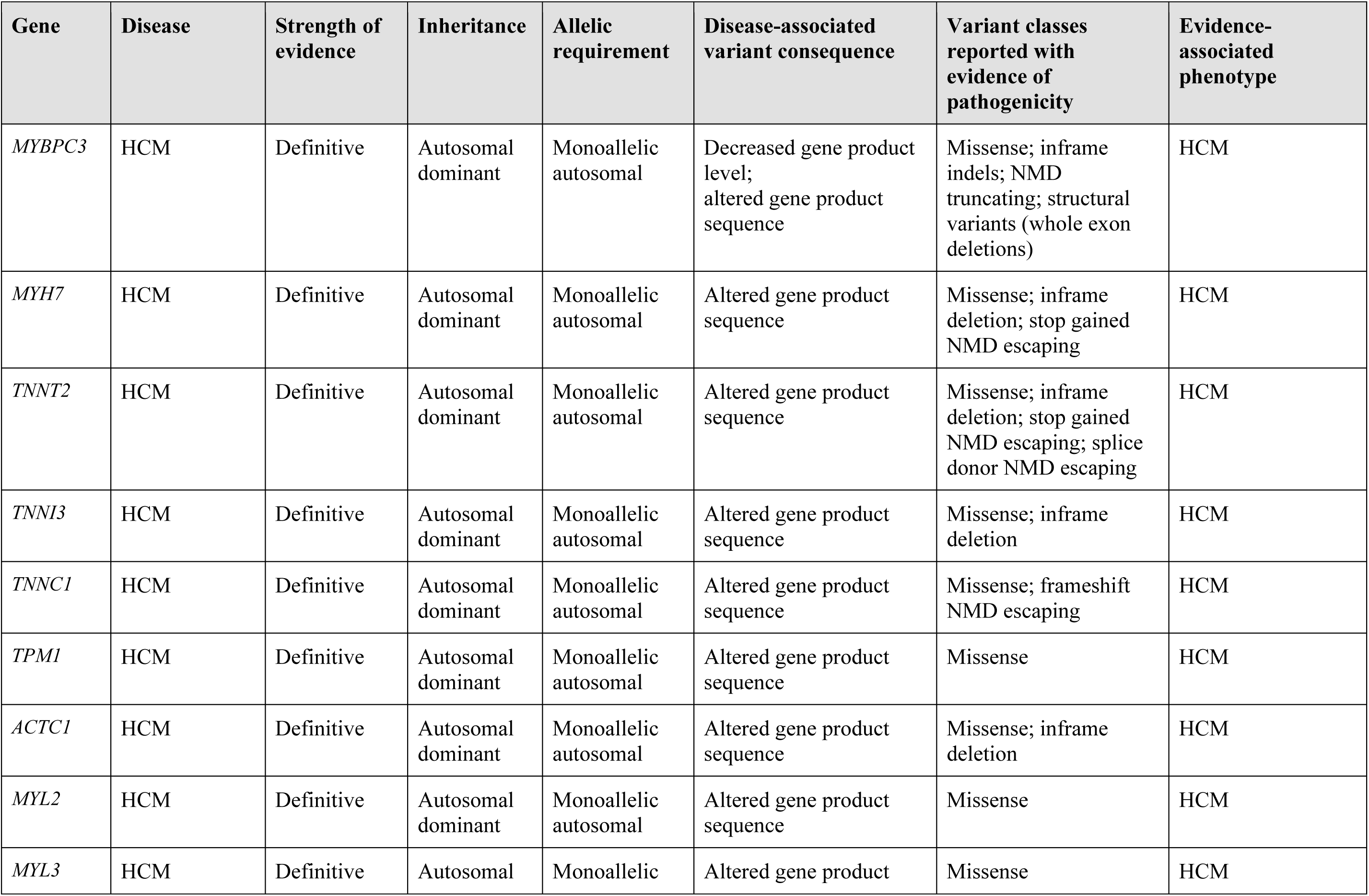
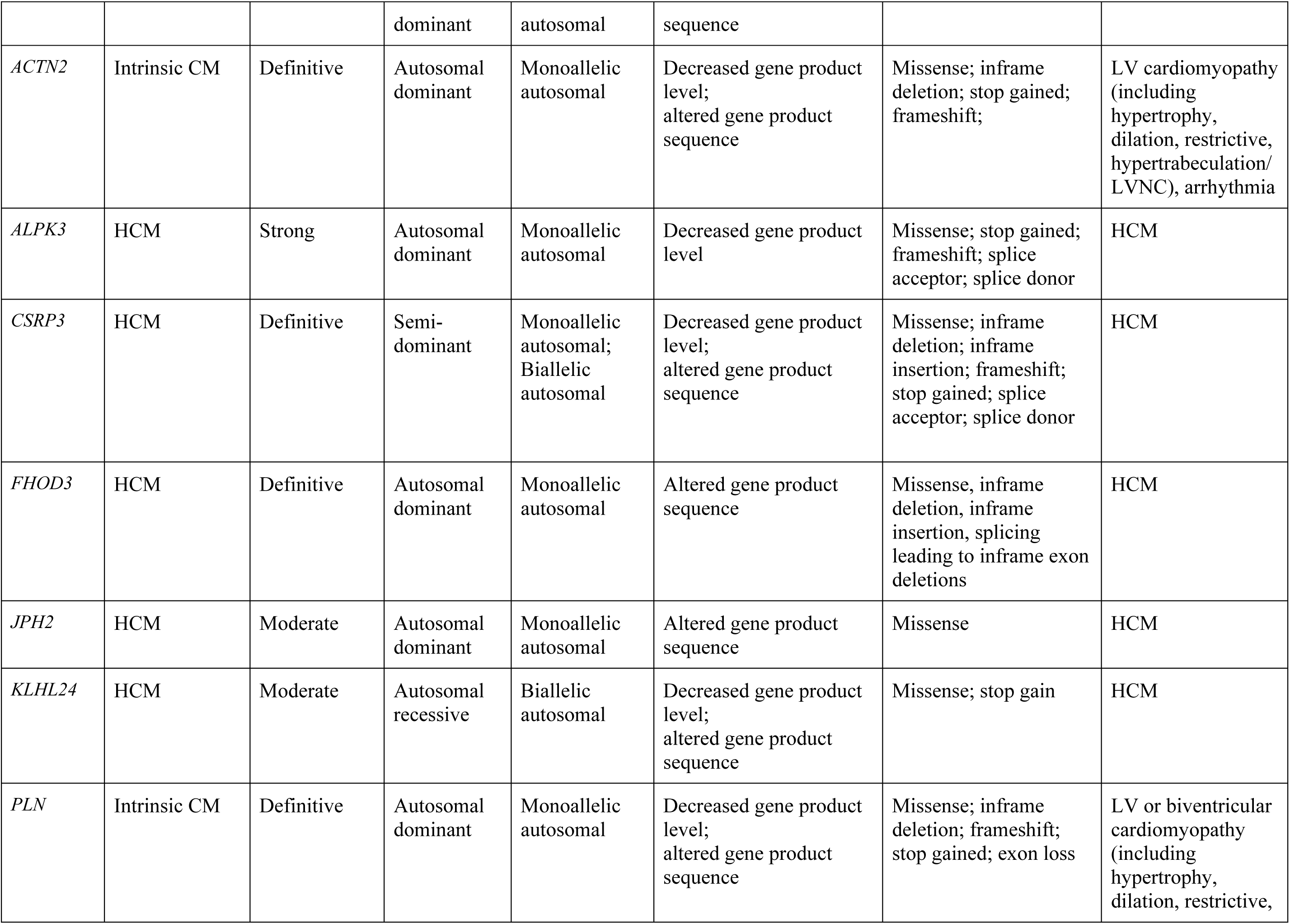
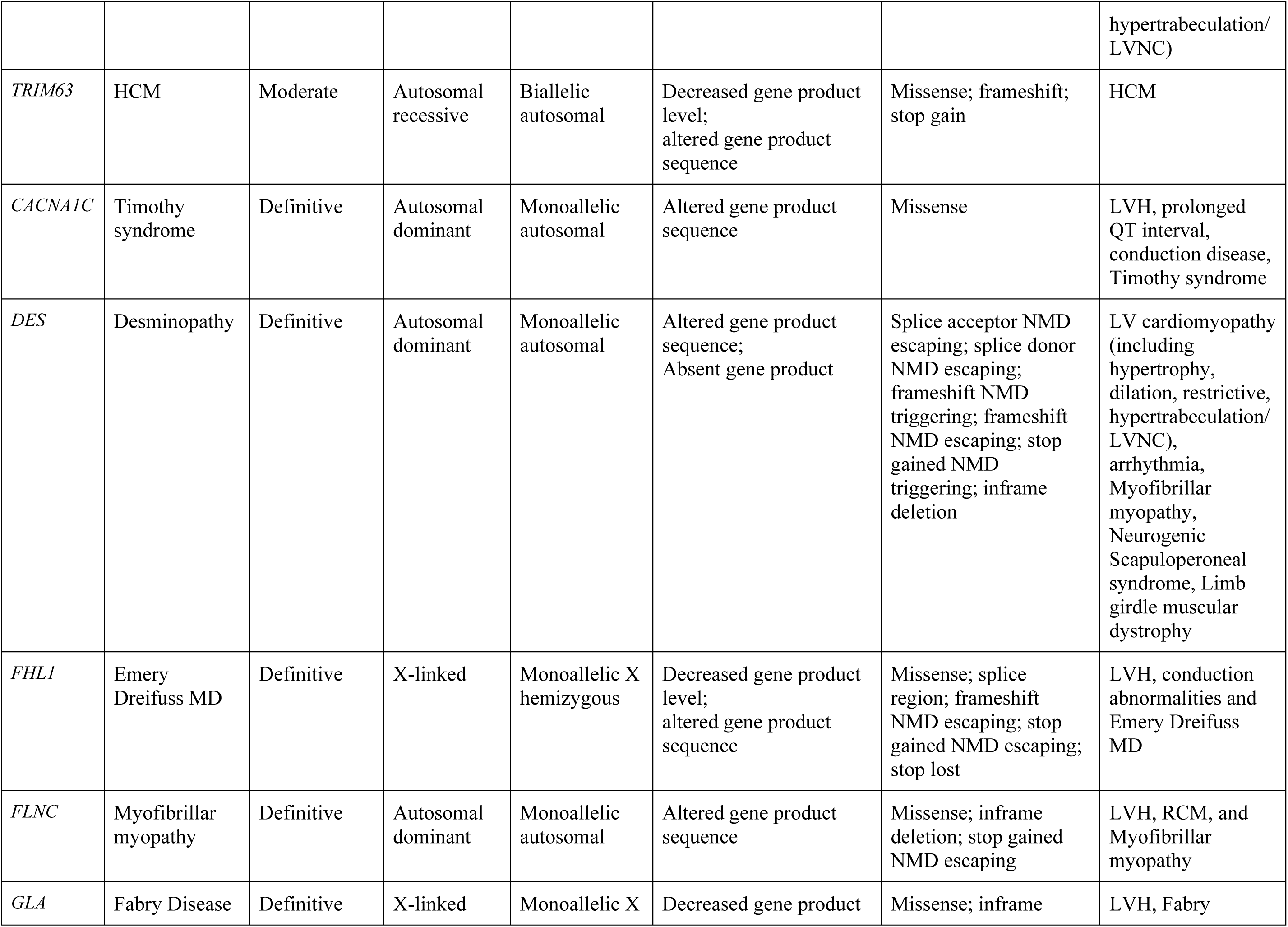
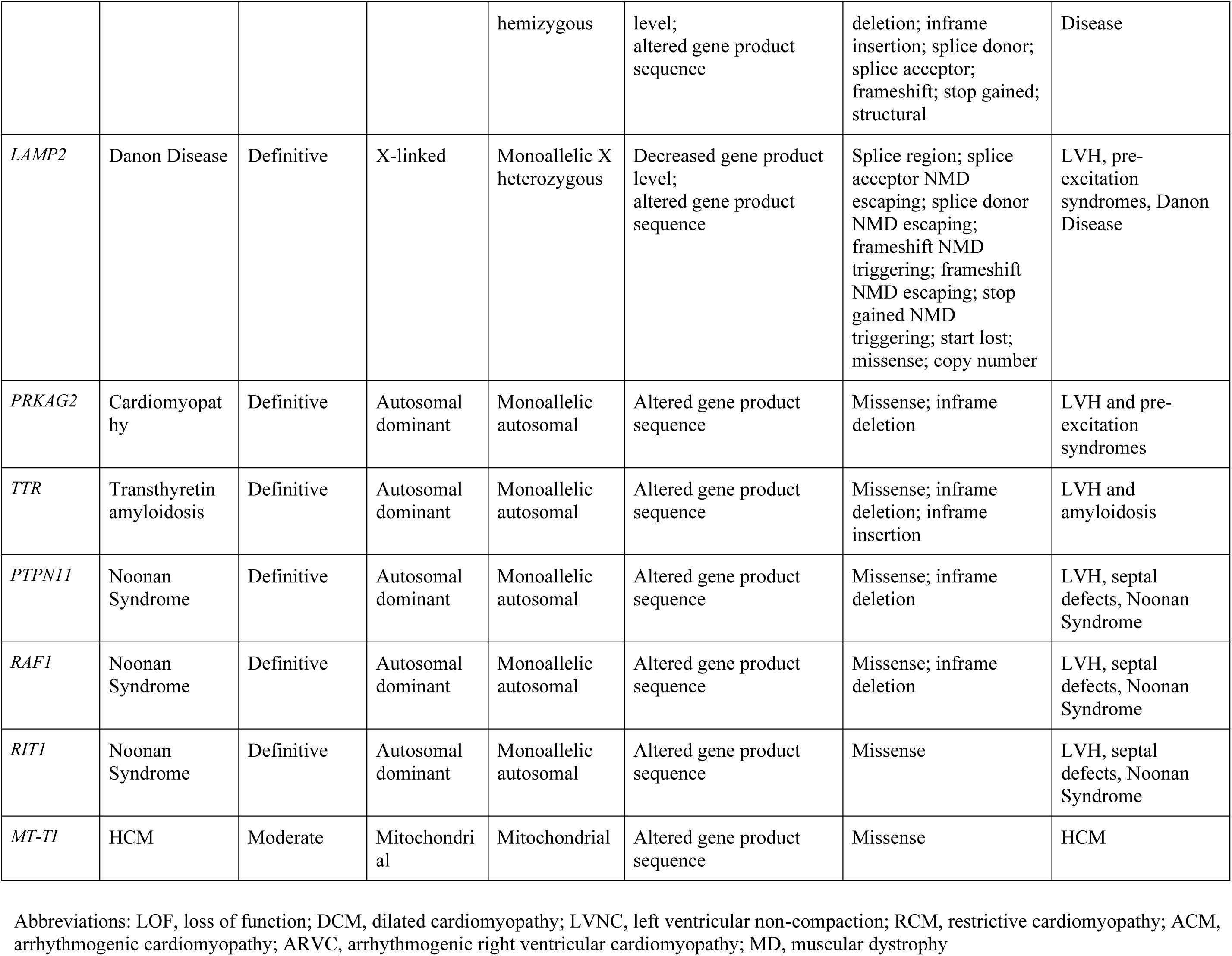
Structured gene-level data for 29 hypertrophic cardiomyopathy (HCM) and syndromic left ventricular hypertrophy (LVH) associated genes.

#### ACTN2

Re-curation of *ACTN2* resulted in an increase from moderate to definitive classification for intrinsic cardiomyopathy. Variants in *ACTN2* have been reported in patients with HCM, dilated cardiomyopathy, LV noncompaction and restrictive cardiomyopathy. This has been repeatedly demonstrated in both the research and clinical diagnostic settings and has been upheld over time. We found no evidence to suggest different molecular mechanisms underlying the disease entities, and indeed individuals can show variable combinations of hypertrophy, dilation, and hyper or hypo-contractility, and the phenotype can vary between individuals in the same family. Based on curated literature, this gene can cause isolated LVH. At least 30 unique heterozygous variants (22 missense, 4 nonsense, 3 deletions, 1 frameshift) and one nonsense homozygous variant have been reported. A majority of the points for this curation came from null variants which often present with hyper-trabeculation in addition to myocardial dysfunction, and indeed truncating variants are enriched in individuals with LV noncompaction compared to controls (0.65% vs 0.01%, p=1.3E-06).^19^

#### CSRP3

Re-curation of *CSRP3* resulted in an increase from moderate to definitive classification, largely due to the addition of a new case-level and experimental data and the re-consideration of a semi-dominant mode of inheritance which allowed for inclusion of recessive case-level data. Semi-dominance includes both dominant and recessive inheritance patterns where individuals with one heterozygous variant have an intermediate phenotype and individuals with a compound heterozygous or homozygous variant have a more severe phenotype and/or earlier onset. Twenty-five unique variants (16 missense, 4 frameshift, 2 nonsense, 2 canonical splice site, and 1 indel) were scored for this curation. Experimental evidence likewise supported this classification, with mouse, cell culture and rescue models^20–22^ and the relationship has been upheld over time.

#### TNNC1

Re-curation of *TNNC1* led to upgrade from moderate to definitive classification for HCM. Variants in *TNNC1* were first reported in HCM patients in 2001.^23^ At least seven papers with genetic evidence supporting the relationship between *TNNC1* and HCM have been published since the original curation of this gene, which allowed for additional genetic evidence to be included in the re-curation. At least eleven unique heterozygous variants (10 missense, 1 truncating) with some evidence to support their pathogenicity and segregation in three families have been reported. Of note, p.Asn144Asp, initially reported in a Belgian cohort,^24^ has now been seen in 23 HCM patients and segregated with disease in 12 affected relatives (personal communication, Tomas Robyns UZ Leuven). *TNNC1* is now the ninth definitive HCM-associated sarcomere gene (in addition to *MYH7, MYBPC3, TNNT2, TNNI3, TPM1, MYL2, MYL3, ACTC1*).

#### ALPK3

New evidence asserting autosomal dominantly inherited loss of function variants in *ALPK3* prompted pre-curation to determine the disease entities to be curated, with a decision to consider this a disease entity distinct from childhood-onset autosomal recessive *ALPK3* cardiomyopathy. At least 38 heterozygous variants (nonsense, frameshift, and splice site) have been reported in probands in at least 5 publications and segregated in ∼ 3 families. Experimental evidence included biochemical function, expression, and models, reaching the maximum score of 6 with the inclusion of recent mouse model and induced pluripotent stem cell functional evidence.^25^ We classified autosomal dominant heterozygous loss of function variants in *ALPK3* as strong for HCM. It is estimated that these variants could account for ∼1% of HCM, though penetrance might be substantially lower than most typical HCM genes. Further detail and real world nuance about this curation is reported in separately.^26^

#### FHOD3

*FHOD3* is a newly curated gene and is classified as definitive for HCM. It was first reported in relation to autosomal dominant HCM in 2018.^27^ Over 50 variants (missense, nonsense, splice variants and in frame del/dup variants) have been reported in >100 probands in at least 6 publications. The majority of variants are located at exon 12 and exon 15 of the cardiac-specific transcript NM_001281740.3, including two recurring variants p.Ser527del and p.Tyr528Cys in exon 12. Most reported variants are missense or in-frame deletions. The gene-disease relationship is supported by expression studies, protein interaction, functional alteration and animal models. However, it should be noted that while experimental studies have shown an important role for *FHOD3* in heart and sarcomeric development, no studies have demonstrated a clear HCM phenotype.

#### TRIM63

We classify *TRIM63* as having moderate evidence of association with autosomal recessive HCM, and disputed for autosomal dominant HCM. *TRIM63* was first reported in relation to autosomal dominant HCM in 2012^28^ with few subsequent clinical reports. Curation of *TRIM63* for autosomal recessive HCM reached moderate classification, and was associated with disease in 32 probands across 5 publications. The Q247X variant is a recurrent variant reported in 11 of 32 probands in either a homozygous or compound heterozygous state. At least 2 reports include co-occurring HCM and skeletal myopathy in the presence of biallelic *TRIM63* variants,^29,30^ though more evidence is needed to understand whether this should be considered part of an expanded phenotypic spectrum. Notably, among multiple reports of autosomal recessive inheritance, none have demonstrated disease in heterozygote relatives.

#### KLHL24

We classified *KLHL24* as moderate for autosomal recessive HCM. Variants were first reported in autosomal recessive HCM in 2019.^31^ Two nonsense and a missense variant have been reported in four probands in three publications. All probands are reported to be from Middle-Eastern countries and consanguineous families. The genetic evidence for these cases was down-scored to avoid over-inflating evidence since it is unlikely that the two alleles arose independently in consanguineous families.

#### MT-TI

*MT-TI* is a mitochondrial gene, which was newly curated and classified as moderate evidence for HCM association. *MT-TI* encodes for mitochondrially-encoded tRNA for isoleucine, it is located on the heavy strand of mitochondrial DNA spanning 69 nucleotides 4263-4331. *MT-TI* was first reported in association with HCM in 1996.^32^ Three variants have been seen in 5 probands in at least 3 publications. One homoplasmic variant m.4300A>G is seen in the majority of cases, including in a family segregating to 16 affected family members.

### Downgraded and notable gene re-curations

Thirteen genes had their classification downgraded to disputed due to a lack of any convincing new evidence in the literature since their initial curation. Genes associated with disease previously (classified as moderate, strong, or definitive) all retained clinically significant classifications after re-curation, whereas genes of uncertain significance (previously classified as limited, disputed, or no known disease relationship) either retained their previous classification or were further downgraded. Clinical laboratories are discouraged from reporting variants in genes with disputed HCM-association.^33^

Of note, evidence for *TTN* variants causing HCM remained limited, rather than being reclassified as disputed. *TTN* has definitive gene-disease validity for dilated cardiomyopathy, but in addition, many variants in *TTN* have been reported in individuals with HCM though the majority are missense without functional data or located in an exon with low percent spliced in (PSI) cardiac tissue. No excess *TTN* variants were noted in cases compared to controls in two studies.^34,35^ There is one report of a frameshift located in the A-band (percent spliced-in; PSI 100%),^36^ and one termination in the I-band (PSI 100%)^37^ in individuals with HCM. One study reported a missense variant in an immunoglobulin domain near the M-line A-band transition zone of *TTN* in a medaka mutagenesis fish model with diastolic dysfunction, highlighting this as an important region. In addition, they reported two missense variants in similar immunoglobulin domains identified in patients with HCM, and showed functional evidence supporting these variants increase MURF1 binding, proposing this as a mechanism for HCM.^38^ Our limited classification acknowledges the need for more research to increase clarity regarding this association.

### Classification updates for syndromic genes

Three genes were selected for re-curation for HCM-like phenotypes that could present as isolated LVH (*CACNA1C*, *FLNC*, *PRKAG2*). While originally classified as definitive, these genes were subsequently re-curated to reflect the updated disease entities they are associated with based on location of variants, variant type, and new literature reports. All retained definitive classifications. Although *CACNA1C* retained a definitive classification for the disease entity of Timothy Syndrome, a specific amino acid position (p.Arg518Cys/His) is reported in association with an isolated cardiac phenotype of LQTS/HCM;^39^ however, current evidence is lacking to confidently split this disease entity from Timothy Syndrome, but will be considered in future. *FLNC* was curated for non-loss of function variants only, after pre-curation supported these variant types as distinct from the arrhythmogenic cardiomyopathy disease entity associated with loss of function *FLNC* variants. Non-loss of function *FLNC* variants were definitively associated with myofibrillar myopathy, with patients presenting with isolated or combined skeletal and/or cardiac features. One variant (p.Trp2710Ter; in last exon) was seen in 36 affected individuals across 6 Hong Kong Chinese families and represents a founder effect. Of note, *FLNC* has a pseudogene located 53.6kb downstream from the functional *FLNC* gene, and exons 46, 47, and 48 are 98% homologous in the functional and pseudogene.^40^ *PRKAG*2 is a gene with overwhelming genetic and experimental evidence supporting a definitive association with glycogen storage disease, however the additional reported phenotype of skeletal myopathy with elevated creatine phosphokinase was included.^41,42^

## DISCUSSION

We report a systematic reappraisal of gene classifications for HCM and syndromes with isolated LVH, including re-curation of genes previously reported by the ClinGen HCM Gene Curation Expert Panel in 2019^16^ and proposed new genes. While the genetic architecture of HCM may be reasonably well understood, there were important updates reflecting our evolving knowledge in the 5 years since the initial HCM gene curation. In total, we identify 29 genes associated with disease, including those with definitive, strong or moderate gene-disease evidence for HCM or isolated LVH. This includes 9 sarcomere genes, with *TNNC1* now considered a definitive evidence gene. In addition, a number of genes, most with sarcomere-associated roles, are associated with HCM. New evidence informed inclusion of *FHOD3* (definitive) and *KLHL24* (moderate), and updated inheritance patterns for *TRIM63* (autosomal recessive; moderate), *CSRP3* (semi-dominant; definitive) and *ALPK3* (autosomal dominant, loss of function variants). Evidence for mitochondrial gene *MT-TI* was evaluated and considered moderate. Nine genes previously considered to have limited or no known disease association, were subsequently further downgraded to disputed gene-disease association, discouraging clinical reporting of variants for HCM patients. There is now greater certainty in the clinical validity of HCM genes following this reappraisal, reflected in the increase of definitive/disputed genes, with fewer genes considered limited or moderate. Additionally, we show the re-curation process did not downgrade any previously disease associated genes, i.e., moderate, strong, and definitive. Systematic reappraisal of disease-associated genes is essential for incorporating new and emerging evidence, and guide appropriate reporting of variants.

HCM has long been considered a genetically heterogeneous disease, though largely due to variants in genes encoding cardiac sarcomere proteins. Genetic testing is a Class I recommendation in all disease guidelines,^1,2,4,7^ and despite decades of research the yield of identifying a likely pathogenic or pathogenic variant in a proband is ∼40%.^43^ Intense research efforts have focused on explaining the remaining genetic causes, with the goal to give greater clarity to patients and their families. In recent years, a sarcomere-negative HCM group has been described, accounting for ∼40% and often being older, more likely male, comorbid hypertension and less severe hypertrophy compared to sarcomere-positive patients.^44,45^ Importantly, it is increasingly clear sarcomere-negative HCM patients likely have a polygenic basis for their disease.^46^ There is now greater recognition of the contribution of genocopies in explaining disease among HCM patients.^10^ Genocopies are described as monogenic conditions that mimic another disease, but have a different genetic etiology, e.g., *TTR*, *PRKAG2* and *LAMP2*. In our previous HCM Gene Curation Expert Panel work, we reflected the nuances of non-sarcomere gene contributions by considering them as syndromic genes, either leading to LVH with or without overt features that could plausibly be misdiagnosed as HCM. This time, our goal was to only curate those genes that could lead to phenotypes that could be mistaken for HCM, i.e., those genes that should be considered for any patient with apparent HCM.

Importantly, genocopies often have additional extra-cardiac features and management considerations, meaning prompt diagnosis is ideal. Indeed, as more targeted therapies for HCM are developed, trialed and incorporated into clinical practice, how these perform is likely to be influenced by the underlying disease etiology. We propose broad recognition of the genetic subtypes that explain apparent HCM, with these including sarcomeric HCM, sarcomere-associated and other monogenic HCM, syndromic LVH (which often include LVH as part of wider phenotypic spectrum of disease and can be mistaken for HCM), and polygenic HCM (Figure 3). Careful clinical and family history, in many cases, will raise the suspicion of the underlying genetic cause of the disease, such as muscle weakness associated with *FLNC* missense variants causing myofibrillar myopathy, or reported consanguinity increasing the likelihood of recessive forms of HCM. We recognize the technical challenges of including a mitochondrial gene in our core HCM gene list, and suggest testing of this gene be considered where the inheritance pattern supports mitochondrial inheritance and the patient is gene-elusive for other HCM-associated genes.

**FIGURE 3:**
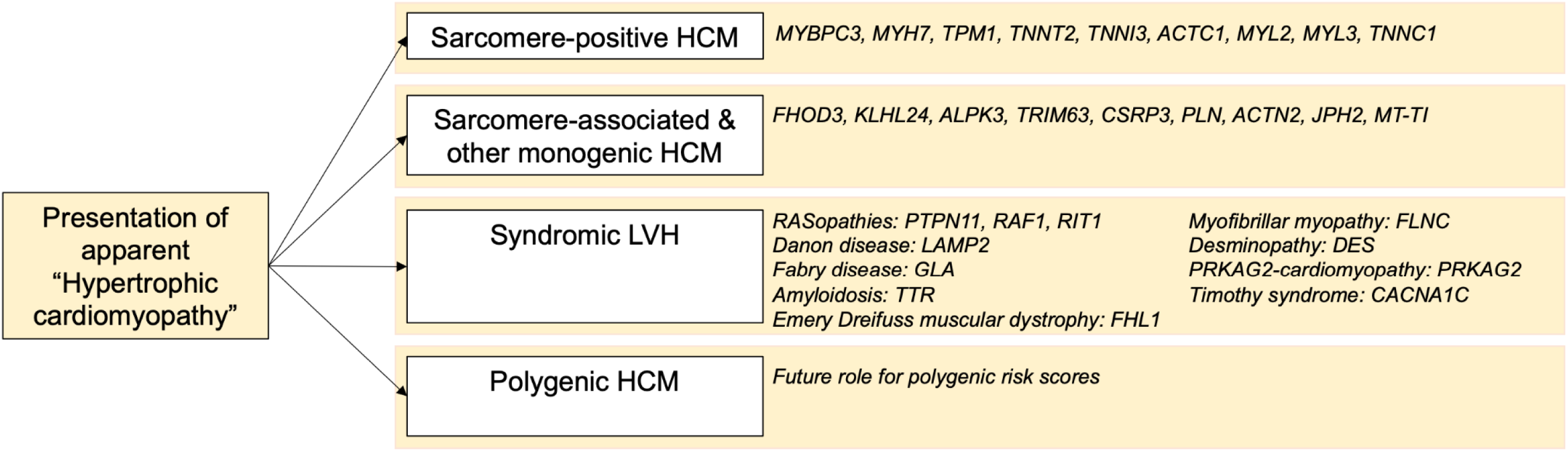
Overview of genetic sub-types of hypertrophic cardiomyopathy and associated genes with moderate, strong or definitive evidence.

Public access to genetic evidence from clinical and research groups globally were key to informing updates to HCM genes. Critically, data that informed some re-classifications were derived from published research studies describing previously underrepresented ancestry groups. In particular, populations with high rates of consanguinity allowed clarification of autosomal recessive and semi-dominant inheritance modes. This includes autosomal recessive gene, *KLHL24* with large Iranian and Saudi Arabian consanguineous families with HCM, sudden cardiac death and cardiac transplant, and accumulation of desmin intermediate filaments in cardiac and muscle biopsy.^31,47^ Likewise, *TRIM63* was re-curated according to an autosomal recessive inheritance pattern, based on numerous studies including a recently published description of HCM in Egyptian patients, accounting for 2% of disease in this population.^48^ Case series and reports of interesting (but not necessarily novel) genomic findings were important in contributing to evidence for many genes, including *ALPK3*,^26^ and highlights the importance of this practice which ensures our ability to openly share findings. In addition, the value of large-scale and comprehensive descriptions from clinical genetic testing laboratories was evident. For example, *CSRP3* was previously curated for only autosomal dominant disease, but based on available evidence considered to have a semi-dominant inheritance pattern,^49^ meaning heterozygous variants can have an intermediate phenotype. How we continue to support open sharing of genomic findings, even when they are not novel research findings and especially for poorly represented ancestry groups, remains critically important to progressing our understanding of the genetics of HCM.

The ongoing sustainability of gene curation efforts needs to be considered. Efforts such as those by ClinGen Gene Curation Expert Panels are time consuming and largely driven by volunteer curators and experts. Our re-appraisal of both previously curated and new HCM genes highlights the value of ongoing effort to update and incorporate new evidence, therefore sustainable models should be considered. In contrast to ClinGen gene curation, other efforts such as PanelApp provide faster, but less in-depth gene-disease associations, using a traffic light system to indicate genes that should or should not be included.^50,51^ Both formats of gene curation are important, and websites that harmonize these various efforts, such as the gene curation coalition (https://thegencc.org/) have become core to genetic test interpretation.^52,53^ Furthermore, we have created a pathway for the community to notify the HCVD Gene Curation Expert Panel of new evidence or requests for genes to be added to curation lists via a link (https://docs.google.com/forms/d/e/1FAIpQLScx9Dl77ScetOqXVLR5oVoxRn3izNdIgTeBGhYkx9tgtlqsjw/viewform?usp=sf_link), in an effort to engage clinical laboratories, clinicians, researchers and consumers in the process. It is reassuring that based on the first re-curation of our initial HCM curation, most genes are now either disease associated (definitive, strong, moderate) or disputed. Ongoing re-curation efforts therefore would target just limited or moderate genes, new reported genes and genes where evidence points to a different phenotype spectrum, inheritance pattern or change in classification, leading to a reduced effort for each subsequent re-appraisal.

Our gene classifications serve as a reflection of our understanding and knowledge at a point in time. Cardiac genomics is a constantly evolving field and efforts to continue to re-appraise gene-disease validity will remain important. We hope that research groups will recognize the framework used for numerically scoring genetic and experimental evidence, and use this as a guide to strengthen the way they report novel gene associations. The ClinGen gene-disease validity curation framework supports curation of monogenic diseases, so more complex inheritance patterns, if applicable, were not included in this analysis. Selection of the gene curation list and the ClinGen gene-disease validity curation framework are both detailed in the Supplemental Data.

## CONCLUSION

While previously considered to be predominantly a disease of the cardiac sarcomere, we report 29 genes with moderate, strong or definitive gene-disease association that should be included in HCM genetic testing. Periodic re-appraisal, including re-curation of known genes and curation of newly asserted genes, is required to incorporate new knowledge and evidence regarding the genetic architecture of HCM.

## Data Availability

All data are publicly available on the ClinGen public repository.

https://clinicalgenome.org/working-groups/clinical-domain/cardiovascular/

## ACKNOWLEDGEMENTS/FUNDING

This publication was supported in part by the National Human Genome Research Institute of the National Institutes of Health through the following grant U24HG009650. JI is the recipient of a National Heart Foundation of Australia Future Leader Fellowship (#106732). BC is a senior clinical investigator of the Research Foundation-Flanders. AB is supported by the 2022 Research Fellowship for aspiring electrophysiologists from the Swiss Heart Rhythm Foundation, and a postdoctoral research fellowship grant from the Gottfried und Julia Bangerter-Rhyner-Stiftung (Switzerland). EMM is supported by NIH HL128075, HG011169, the Leducq Foundation, and the American Heart Association. JW is supported by Medical Research Council (UK), Sir Jules Thorn Charitable Trust [21JTA], British Heart Foundation [RE/18/4/34215], and the NIHR Imperial College Biomedical Research Centre.

